# The Prevalence and Factors Associated with Prompt Diagnosis and Treatment of Fever among Under-five Children in Zambia; Evidence from a Country-wide Cross-Sectional Survey

**DOI:** 10.1101/2023.09.05.23295058

**Authors:** Khumbutso Elizabeth Phiri, Cephas Sialubanje, Busiku Hamainza, Japhet Chiwaula, Mukumbuta Nawa

## Abstract

**Introduction:** Fever is one of the signs of a suspected infection as the body mounts an inflammatory response to fight infections such as malaria, pneumonia and gastroenteritis. Prompt diagnosis of the aetiology of the fever and appropriate treatment can prevent severe disease and mortality. Delayed diagnosis and treatment of infections contribute significantly to the high under-five mortality ratio. This study assessed the prevalence and associated factors to prompt diagnosis and treatment of fever among under-five children in Zambia.

**Methods:** The study used secondary data from a nationwide cross-sectional survey carried out during the rainy season, a peak transmission season for malaria and oral-faecal transmitted diseases. The survey used multistage computer-generated random sampling by statistical enumeration areas (SEAs) as clusters and households within the SEAs. Informed consent was obtained from the adult caregivers of the children. All children within the selected households below five years were included in the study, and the caregivers were interviewed on the occurrence of fever in the two weeks preceding the survey. In addition, the children were tested for malaria using rapid diagnostic tests and haemoglobin count. The data was analysed in STATA version 14 using weights to account for inter and intra-cluster correlations (ICC). Descriptive statistics using measures of frequencies, medians and interquartile ranges were done, and cross-tabulations and logistic regression were used to assess measures of association. The significance level was set at a P-value of 0.05 and a confidence level of 95%.

**Results:** Out of the 3003 under-five children included, 728 had a fever in the two weeks preceding the survey, reflecting the prevalence of fever of 19.5%. The fever was more common in poverty-related indicators such as lower education levels of heads of households, rural areas, lower wealth status, not using ITNs, and malaria and anaemia. The prevalence of prompt health-seeking behaviour was 57.3% which raises concerns that 42.7% of the children with fever did not have prompt treatment within the same of the next day of fever onset. Cross-tabulations showed that prompt health-seeking behaviour was more common in children who were boys, children older than one year compared to those who were below one year, heads of households with secondary or tertiary education, Eastern, Muchinga and North-western provinces and those who did use indoor residual spraying. Further, the adjusted multivariable model found that the male sex of the children was associated with significantly increased odds of prompt health-seeking behaviour aOR 1.52 and the use of IRS was also associated with increased odds of prompt health-seeking behaviour aOR 1.8. While having secondary or tertiary education compared to no education was not statistically associated with prompt health-seeking behaviour (P-value = 0.085), the odds were also increased nonetheless aOR 1.30.

**Conclusion:** This study has found that the prevalence of fever among under-five children in Zambia is comparable to other sub-Saharan African Countries. Fever was associated with age, education, area of residence, wealth status and use of malaria prevention interventions like insecticide-treated nets and indoor residual spraying. Further, fever was found to be associated with the presence of anaemia and malaria. Health-seeking behaviour among children with fever was sub-optimal, as a considerable proportion did not seek treatment. Factors associated with health-seeking behaviour among children with fever include sex, education levels of the head of the household and staying in a house with indoor residual spraying.

## Introduction

Fever is a common symptom of various illnesses, especially among children, and is often a sign of an inflammatory response or an underlying infection such as malaria (Mutanda, Cheruiyot et al. 2014). Timely diagnosis and appropriate treatment are crucial in managing febrile illnesses, as delays can lead to complications and worsened outcomes (Nawa, 2020). Children, especially those under the age of five, are particularly vulnerable to the adverse effects of fever due to their developing immune systems. Untreated fevers can become serious illnesses like bacterial infections, viral diseases, and severe malaria (Basu & Sahi, 2017). Prompt diagnosis and appropriate treatment reduce the risk of complications, alleviate parental anxiety, and improve the child’s quality of life (Basu & Sahi, 2017).

One of the primary determinants of prompt diagnosis and treatment of fevers, including malaria infection, is the accessibility and quality of healthcare services (Nawa, 2019). Inadequate access to healthcare facilities, particularly in rural and remote areas, can lead to delays in seeking medical attention (Nawa, Halwindi et al. 2020). Distance, transportation availability, and healthcare infrastructure are pivotal in these delays (Sasaki, Fujino et al. 2010; Menda, Nawa et al. 2021).

Overburdened healthcare systems and long waiting times can discourage parents from seeking immediate care for their febrile children (Stekelenburg, Kyanamina et al. 2004). Socioeconomic status significantly influences a family’s ability to seek prompt medical care for a child with a fever (Danny, Nawa et al., 2023). Families with lower income levels may face financial barriers that prevent them from accessing healthcare services promptly (Sialubanje, Massar et al. 2015). Whilst health primary health care services in Zambia are free, associated costs, such as transport costs, diagnostic tests, and medications, can deter parents from seeking early medical attention and prioritise immediate needs such as food (Chanda-Kapata, Kapata et al., 2016).

Parental knowledge and awareness of fever-related illnesses and their potential complications are crucial in determining how promptly medical care is sought (Fujino, Sasaki et al., 2009). Educated parents are more likely to recognise the seriousness of fever and seek medical attention earlier (Fujino, Sasaki et al., 2009). Conversely, a lack of understanding about the significance of fever, its potential causes, and appropriate treatment options can result in delayed healthcare-seeking behaviour (Kaiser, Fong et al., 2019). Cultural beliefs and practices within communities can significantly influence healthcare-seeking behaviours. Some cultures may prioritise traditional remedies or alternative therapies over conventional medical interventions (Maritim, Silumbwe et al. 2021). This can lead to delays in seeking professional medical care, allowing the underlying illness to progress. Bridging the gap between traditional beliefs and modern healthcare practices through culturally sensitive education can encourage families to seek prompt diagnosis and treatment (Sivalogan, Banda et al., 2023). Other factors include language barriers, fear of medical procedures, and distrust in healthcare providers, all contributing to delays in seeking care (Mwamba, Beres et al., 2022).

This study aimed to find the prevalence of prompt health-seeking behaviour among children with fever and associated factors. This will help program managers and policymakers to understand why children continue to bear the highest burden of morbidity and mortality of treatable fever-associated diseases such as malaria in Zambia.

## Methodology

### Study Design

This is a secondary analysis of data collected during a survey called the Malaria Indicator Survey.

### Study Settings

The study covered all the ten provinces of Zambia. Zambia has a tropical climate with high temperatures and annual rainfall, especially in the country’s north. Savannah grasslands, river valleys and forested plateau land cover it. The climate and landscape are conducive to breeding and proliferating malaria mosquito vectors.

### Sample Size

The main study was powered to detect malaria prevalence in Zambia at a 95% confidence level and power of 80% at the provincial level and urban versus rural strata. It was not designed to detect fever prevalence. Based on the 41% prevalence of fever detected in the Zambia Demographic and Health survey of 2018, we used the survey sample size.

n = Z^2^ p(1-p)/d^2^ where Z = 1.96 at 95% confidence level, p = 0.41 and d = 0.05

= 1.96 × 0.41x0.59/(0.05x0.5) = 371

Further, a 1.5 inflation factor was used to account for multi-cluster sampling and non-response to a minimum of 557.

### Sampling Procedure

We included all children in the primary study who met our criteria based on age six to 59 months and who had slept under a net the night before the survey. The survey used multistage computer-generated random sampling by statistical enumeration areas (SEAs) as clusters and households within the SEAs. All children aged six to 59 months within selected households were included in the study.

### Data Collection

We extracted the data from the Malaria Indicator Survey database at the National Malaria Elimination Centre into Microsoft Excel and then imported it into Stata for cleaning and analysis. The primary survey used a questionnaire that trained research assistants administered.

### Data Analysis

The data was analysed in Stata software version 14 using complex data analysis to account for the multistage sampling. We conducted descriptive statistics using counts, percentages, mean or median and confidence intervals depending on the distribution. We did parametric or non-parametric hypothesis tests to compare among sample groups. We used logistic regression using stepwise forward model building for associations by conducting univariate and multivariable analysis. Variables with a P value of 0.1 or less in the univariate model were included in the multivariable regression model. A P value of 0.05 was considered significant, and the Akaike and Bayesian information criteria were used to assess model fitness.

### Ethical Considerations

The data was anonymised with coded identity numbers that were not traceable to the participants. In the primary survey, the heads of the households consented to participate in the study. The study had ethical clearance from the University of Zambia Biomedical Ethics Committee (UNZABREC).

## Results

This study found that 728 children under the age of five had a fever in the two weeks preceding the survey, indicating a prevalence of fever of 19.5% (95%CI 16.6 – 23.4%). Out of these, 57% sought treatment. There is a huge gap of 42.7% of children who run fevers but did not seek treatment and, therefore, run the risk of developing complications such as severe malaria. While the main survey that used the general population of children below the age of five years had reported a malaria prevalence of 39%, our sub-analysis using only children who had a fever in the past two weeks indicated a higher malaria prevalence of 54% (95%CI 49.1 – 58.9%). This indicates that if a child runs a fever, there is more than a 50% likelihood that he or she has malaria. The sex distribution was at par; the median age was 33 months, but there were more older children than those below one year (89.7%). For those who ran a fever, the majority came from rural areas (93%) and came from the lowest socioeconomic status. Over half of the households were sprayed with Indoor Residual Spraying, with the median being about five months before the survey, and 45% used insecticide-treated nets. Over 60% of the children had anaemia; the median haemoglobin count was 10.5 mg/dl. Table 1 summarises the descriptive statistics and estimates of some key variables.

Regarding health-seeking behaviour by various variables, there was a statistically significant difference between male and female children. This finding may be incidental; however, there was a significant difference among children aged below one year and those older than one year. Different education levels of household heads were also statistically different regarding health-seeking behaviour. Those with tertiary and secondary were more likely to seek treatment for their children with fevers than those with no or primary education. Other variables that showed significant differences in health-seeking behaviours for children with fevers included provinces and those whose houses were sprayed with indoor residual chemicals. The variables that showed no statistical differences included where one’s house was located, whether rural or urban, wealth quintiles, use of Insecticide Treated Nets, whether one had anaemia or not and whether one tested positive for malaria using Rapid Diagnostic Tests. Table 2 summarises the findings of the cross-tabulation between health-seeking behaviour and different variables.

Further analysis using logistic regression to measure factors associated with health-seeking behaviour among children aged below five years who had a fever in the two years preceding the survey showed that three variables met the inclusion criteria of P-Value less than 0.2 using the univariate model, namely sex, highest education of the head of the household and living in a house that was sprayed with Indoor Residual chemicals. When the multivariate model was fitted with the three included variables, one variable (highest educational level of the head of the household) became statistically insignificant, whilst sex and IRS remained significant. When the highest educational level of the head of the household was removed, the other two variables, sex and IRS, remained statistically significant. However, the coefficient of determination (pseudo-R^2^) reduced from 0.0275 in the full model to 0.0192 in the reduced model, and the Akaike Information criteria increased from 829.42 to 980.61 whilst the Bayesian Information criteria increased from 847.18 to 994.39 respectively. Based on this model fitness assessment, the three-variable model (sought treatment, sex and IRS) was discarded, and the four-variable model (sought treatment, sex, highest education and IRS) was maintained because it had the lower Akaike and Bayesian Information criteria and a higher coefficient of determination. Table 3 presents the measures of association of health-seeking behaviour among children with fever in this study.

## Discussion

This study set out to determine the prevalence of fever among under-five children in Zambia and assess the factors associated with prompt diagnosis and treatment of children with fever. The findings are that about a fifth (19.5%) of the children had an episode of fever within the two weeks preceding the survey. One study in Zambia found a fever prevalence of 1.5%; however, this was a health facility survey, not a community survey like ours (Chanda, Hamainza et al., 2009). Another study, a household survey, found the prevalence of fever to be 13.4% among under-five children in Zambia (Chirwa, Malinga et al., 2023). This study may have found a lower prevalence because it only had data from 28 districts from eight of the provinces in Zambia, which did not include high malaria prevalence provinces like Luapula. In contrast, our study included all ten provinces. Our study findings are comparable or even lower to studies elsewhere in sub-Saharan Africa, which found the prevalence of fever in under-five children to be 16% in Nigeria, 20% in Ghana, 25% in Kenya and 26% in Sierra Leone (Novignon & Nonvignon, 2012).

Among those with a fever, it was equally common in both male and female children. This is similar to another multi-country study, which found no sex differences in Africa (Baume & Marin, 2007). In the Nonvignon and Nonvignon (2012) study of four sub-Saharan countries, only children from Ghana had shown that there were significant sex differences in the prevalence of fever in under-five children; the other three countries, Nigeria, Kenya and Sierra Leone showed no such sex differences. In the under-five children age group, there is no biological plausibility of unequal exposure to infections among the sexes; however, boys are more prone to injuries than girls, especially among older children (Stracciolini, Casciano et al. 2014). Boys tend to engage in outdoor and adventurous activities such as combative sports (Stracciolini, Casciano et al. 2014). With age, our study found that older children had a higher prevalence of fever than children below one year. This may be because children below one year are more likely to be cared for by their caregivers, whilst older children may receive less care, such as warm clothing, sleeping under insecticide-treated nets and receiving full vaccinations (Chard, Gacic-Dobo et al. 2020). Fever prevalence was also higher in those in the lowest wealth bracket, rural areas and those from households whose heads had no or only primary education. Other studies supported these findings (Uzochukwu, Onwujekwe et al., 2008; Weisz, Meuli et al., 2011). Children from wealthier families, urban areas and educated caregivers are likely to receive better nutrition and care, full vaccination and other prevention interventions such as insecticide-treated nets, better housing and water sources (Novignon & Nonvignon, 2012).

Further, fever was more common in those who did not use preventive interventions like indoor residual spraying and insecticide-treated nets than those who used them. It was also more common in those who tested positive for malaria than those who tested negative. This implies that malaria contributes significantly to the occurrence of fever in Zambia. However, as can be seen, not all fevers were malaria; some with no malaria evidence still had fevers. Our study was corroborated by other studies elsewhere in Africa and Asia (Kumar, Chery et al. 2012; Prasad, Sharples et al. 2015; Dalrymple, Cameron et al. 2017). Fever was also more common in those who were anaemic than those who were not. Our observational study does not elicit causal relationships. However, this may be due to a dual role-play where those with lower blood indices have weaker immunities to infections, and those with infections have haemolysis and show lower blood indices such as haemoglobin.

Almost two-thirds (57.3%) of the children who had experienced fevers sought treatment from health facilities. However, It is worrying that a good proportion (42.7%) did not seek treatment because lack of or delayed treatment of febrile childhood illnesses can lead to severe disease manifestation and mortality within very short periods (Hildenwall, Tomson et al. 2008). Cross-tabulations of associated factors for seeking treatment using the chi-square test of association and Fisher’s exact test indicated that sex, age, education levels, province and indoor residual spraying were significant. In contrast, area of residence (rural versus urban), wealth status, ITN use, malaria status and anaemia were not significant factors in seeking treatment among children with fever. Further analysis using logistic regression to adjust for confounding revealed that only sex and use of indoor residual spraying were significantly associated with seeking treatment among children with fevers. This analysis demonstrates that merely running descriptive statistics and cross-tabulations with hypothesis tests in observational studies may lead to making false observations and conclusions due to confounding if the analyses are not adjusted for confounders (Tolles and Meurer 2016, Schuster, Twisk et al. 2021). The education level of the household head was found to be statistically significant in the bivariate regression, but in the multivariable analysis, it became insignificant. The main multivariable model with sex, education level of the household head and the use of IRS had a higher coefficient of determination (pseudo-R^2^) and lower Akaike and Bayesian Information Criteria than the model where educational level was removed. This indicates that though the educational level was not statistically significant, it was still an important predictor in improving model fitness. The higher odds ratio concerning the base (no or primary education) indicates that more education is associated with better outcomes in health-seeking behaviour. The higher educational level of the household head has been found in other studies to be an important predictor of health-seeking behaviour, as educated caregivers are more likely to identify danger signs, make quicker decisions, and have the means and access to health services (Adedokun & Yaya, 2020).

The sex of the child was found to be a significant predictor of health-seeking behaviour. African societies, including in Zambia especially in rural areas, are patriarchal and consider male children more as they carry the family name and continue with family legacies, whilst female children are married off and bear the names of their husbands (Ogundipe-Leslie, 1993; Dyer, 2007). On the other hand, some African cultures value women more as they enhance family wealth when the family charges the bride price at the time of marriage (Taringa & Museka, 2021). In this study, we do not know whether these cultural factors influenced our findings on why male children were associated with better health-seeking behaviour compared to female children, as this was an observational study with no qualitative explanatory inquiry. Indoor Residual spraying was also significant in our study; we did not find a direct association between deployment of IRS and health-seeking behaviour. The indirect relationship is that those who accessed the IRS were also more likely to access other interventions, including access to health services (Hailu, Lindtjørn et al. 2016).

## Conclusion

This study has found that the prevalence of fever among under-five children in Zambia is comparable to other sub-Saharan African Countries. Fever was found to be associated with age, education, area of residence, wealth status and use of malaria prevention interventions like insecticide-treated nets and indoor residual spraying. Further, fever was found to be associated with the presence of anaemia and malaria. Health-seeking behaviour among children with fever was sub-optimal, as a considerable proportion did not seek treatment. Factors associated with health-seeking behaviour among children with fever include sex, education levels of the head of the household and staying in a house with indoor residual spraying.

### Recommendations

This study recommends that more interventions be given to caregivers of under-five children, such as sensitisation on danger signs such as fever and provision of interventions such as ITNs and IRS. More research is needed in order to understand disparities in health-seeking behaviour based on the child’s sex in order to address gender equity.

### Strengths and Limitations

This study covered the whole country and was adequately powered to represent national, provincial level and urban and rural strata. In addition, the study adjusted for confounding among social demographic factors such as age and sex interventions such as IRS and ITN use; the study further adjusted for biological factors such as anaemia, which can precipitate or be a consequence of infections. This was an observational study, so it could not elicit cause-effects but associated factors so that no causal inferences could be generated. Further, because the study used secondary data, it could only use the available data, and some key factors, such as vaccination status, could not be included.

## Supporting information

Table 1 Summary Statistics

Table 2 Cross-tabulations

Table 3 Factors Associated with Health Seeking Behavior

## Data Availability

All data produced in the present study are available upon reasonable request to the authors

