## Supplementary material for "The Prevalence and Factors Associated with Prompt Diagnosis and Treatment of Fever among Under-five Children in Zambia; Evidence from a Country-wide Cross-Sectional Survey": Table 1 Summary Statistics

**Table 1: Summary of the Descriptive Statistics**

| **Summary of Categorical Variables** | | | |
| --- | --- | --- | --- |
| **Variable** | **Categories** | **Number (n = 728)** | **Percentage (95% CI)*** |
| Sex | Male | 365 | 50.1 (45.0 - 52.2) |
|  | Female | 363 | 49.9 (44.8 - 55.0) |
| Age (Years) | Below 1 Year | 77 | 10.6 (3.6 - 17.3) |
|  | One year | 133 | 18.3 (11.7 - 24.9) |
|  | Two year | 174 | 23.9 (17.6 - 30.2) |
|  | Three Years | 156 | 21.4 (15.0 - 27.8) |
|  | Four Years | 188 | 25.8 (19.5 - 32.1) |
| Education Household Head | No or Primary | 376 | 51.6 (46.5 - 56.7) |
|  | Secondary | 229 | 31.4 (23.4 - 37.4) |
|  | Tertiary | 21 | 2.9 (-4.3 - 10.1) |
| Residence Type | Rural | 677 | 93.0 (92.2 - 95.8) |
|  | Urban | 51 | 7.0 (5.8 - 8.3) |
| Wealth Quintiles | 1 (Lowest) | 218 | 30.0 (23.9 - 36.1) |
|  | 2 (Upper Low) | 148 | 20.3 (13.8 - 26.8) |
|  | 3 (Middle) | 143 | 19.6 (1.1 - 26.1) |
|  | 4 (Lower High) | 156 | 21.4 (15.0 - 27.8) |
|  | 5 (Highest) | 63 | 8.7 (7.9 - 9.5) |
| House Indoor Residual Spray | Sprayed | 420 | 57.7 (53.0 - 62.4) |
|  | Not Sprayed | 308 | 42.3 (36.8 - 47.8 ) |
| ITN Use | Use ITN | 333 | 45.7 (40.3 - 51.1) |
|  | Not Use ITN | 62 | 8.5 (1.6 - 15.4) |
| Anaemia | Anaemic | 457 | 62.8 (58.3 - 67.3) |
|  | Not Anaemic | 251 | 37.2 (31.2 - 43.2) |
| Malaria (RDT) | Positive | 393 | 54.0 (49.1 - 58.9) |
|  | Negative | 335 | 46.0 (40.7 - 51.3) |
| Ill with Fever | Yes | 728 | 19.5 (16.6 - 23.4) |
|  | No | 3003 | 80.5 (79.1 - 81.9) |
| Sought Treatment | Yes | 417 | 57.3 (52.6 - 62.0) |
|  | No | 311 | 42.7 (37.2 - 48.2) |
| **Summary of Continuous Variables** | | | |
| **Variable** | **Units** | **Median** | **Inter Quartile Range** |
| Age | Months | 33 | (20 - 48) |
| Last IRS | Months | 5 | (5 - 6) |
| Last Fever | Days | 3 | (2 - 7) |
| Haemoglobin | mg/dl | 10.5 | (9.2 - 11.5) |

*A test of proportions was used to calculate the confidence intervals for point estimates
