## Supplementary material for "The Prevalence and Factors Associated with Prompt Diagnosis and Treatment of Fever among Under-five Children in Zambia; Evidence from a Country-wide Cross-Sectional Survey": Table 2 Cross-tabulations

**Table 2 Health-Seeking Behaviour by Different Variables**

|  |  | **Sought Treatment** | | |
| --- | --- | --- | --- | --- |
| **Variable** | **Categories** | **Not Sought Tx n(%)** | **Sought Tx n(%)** | **P-Value*** |
| Sex | Male | 132 (36.4) | 231(63.6) | 0.001 |
|  | Female | 179 (49.0) | 186 (51.0) |  |
| Age (Years) | Below 1 Year | 40 (52.0) | 37 (48.1) | 0.02 |
|  | One year | 45 (33.8) | 88 (66.2) |  |
|  | Two year | 67 (38.5) | 107 (61.5) |  |
|  | Three Years | 78 (50.0) | 78 (50.0) |  |
|  | Four Years | 81 (43.1) | 107 (56.9) |  |
| Education Household Head | No or Primary | 160 (42.6) | 216 (57.5) | 0.034 |
|  | Secondary | 90 (39.3) | 139 (60.7) |  |
|  | Tertiary | 3 (14.3) | 18 (85.7) |  |
| Residence Type | Rural | 286 (42.3) | 391 (57.8) | 0.346 |
|  | Urban | 25 (49.0) | 26 (51.0) |  |
| Province | Central | 13 (40.6) | 19 (59.4) | 0.006 |
|  | Copperbelt | 42 (51.2) | 40 (48.8) |  |
|  | Eastern | 37 (28.9) | 91 (71.1) |  |
|  | Luapula | 114 (48.5) | 121 (51.5) |  |
|  | Lusaka | 11 (64.7) | 6 (35.3) |  |
|  | Muchinga | 17 (30.4) | 39 (69.6) |  |
|  | Northern | 23 (43.4) | 30 (56.6) |  |
|  | North-Western | 20 (40.0) | 30 (60.0) |  |
|  | Southern | 6 (42.9) | 8 (57.1) |  |
|  | Western | 28 (45.9) | 33 (54.1) |  |
| Wealth Quintiles | 1 (Lowest) | 97 (44.5) | 121 (55.5) | 0.8 |
|  | 2 (Upper Low) | 66 (44.7) | 82 (55.4) |  |
|  | 3 (Middle) | 61 (32.7) | 82 (57.3) |  |
|  | 4 (Lower High) | 60 (38.5) | 90 (61.5) |  |
|  | 5 (Highest) | 27 (42.9) | 36 (57.1) |  |
| House Indoor Residual Spray | Sprayed | 162 (38.6) | 258 (61.4) | 0.008 |
|  | Not Sprayed | 149 (48.4) | 159n(51.6) |  |
| ITN Use | Use ITN | 128 (38.4) | 205 (61.6) | 0.218 |
|  | Not Use ITN | 29 (46.8) | 33 (53.2) |  |
| Anaemia | Anaemic | 203 (44.4) | 254 (55.6) | 0.282 |
|  | Not Anaemic | 101 (40.2) | 150 (59.8) |  |
| Malaria (RDT) | Positive | 175 (44.5) | 218 (55.5) | 0.285 |
|  | Negative | 136 (40.6) | 59.4 (59.4) |  |

*Pearson's Chi-Square Test of Association was used to calculate P values except for the education levels of household heads, where some cells were below five, and Fisher's Exact Test was used instead.
