## Supplementary material for "The Prevalence and Factors Associated with Prompt Diagnosis and Treatment of Fever among Under-five Children in Zambia; Evidence from a Country-wide Cross-Sectional Survey": Table 3 Factors Associated with Health Seeking Behavior

**Table 3: Factors Associated with Health-Seeking Behaviour Among Under-Five Children with Fever**

| **Variable** | **Category** | **Unadjusted OR** | **95%CI** | **P-Value** | **Adjusted OR** | **95%CI** | **P-Value** |
| --- | --- | --- | --- | --- | --- | --- | --- |
| Sex | Female | 1 |  |  | 1 |  |  |
|  | Male | 1.68 | 1.25 - 2.27 | 0.001 | **1.52** | **1.10 - 2.12** | **0.011** |
| Age | 0  1- 4 years | 1  0.97 | 0.87-1.08 | 0.557 |  |  |  |
| Residence | Rural | 1 |  |  |  |  |  |
|  | Urban | 0.76 | 0.43 - 1.34 | 0.347 |  |  |  |
| Education | No &Primary  2^o^ & 3^o^ | 1  1.33 | 1.00 - 1.79 | 0.049 | 1  1.3 | 0.97 - 1.74 | 0.085 |
| Province | 10 Provinces | 1 | 0.94 - 1.06 | 0.927 |  |  |  |
| Wealth Quintile | Poorest Q  2 - 5 Q | 1  1.05 | 0.94 - 1.18 | 0.342 |  |  |  |
| IRS | No IRS | 1 |  |  | 1 |  |  |
|  | IRS Yes | 1.49 | 1.11 - 2.01 | 0.008 | **1.8** | **1.30 - 2.50** | **< 0.001** |
| ITN Use | No ITN Use | 1 |  |  |  |  |  |
|  | ITN Use | 0.71 | 0.41 - 1.23 | 0.22 |  |  |  |
| Anaemia | Anaemia (< 11 mg/dl) | 1 |  |  |  |  |  |
|  | No Anaemia (≥ 11 mg/dl) | 1.19 | 0.87 - 1.62 | 0.282 |  |  |  |
| Malaria (RDT) | Positive | 1 |  |  |  |  |  |
|  | Negative | 1.17 | 0.87 - 1.58 | 0.285 |  |  |  |
